# Exploring the Genetic Relationship Between Migraine Subtypes, Depression And Anxiety Using Polygenic Scores

**DOI:** 10.64898/2026.06.21.26355498

**Authors:** A. Christelle Doppenberg, Dale R. Nyholt, Nicholas C. Martin, Naomi R. Wray, Ian Hickie, Catherine M. Olsen, David C. Whiteman, Jodi T. Thomas, Brittany L. Mitchell

## Abstract

Migraine is a disabling neurological disorder that frequently co-occurs with depression and anxiety. While prior research suggests a genetic association between these conditions, little is known about the genetic relationships underlying specific migraine subtypes. Bivariate genetic correlations between migraine with depression and anxiety were estimated using Linkage-Disequilibrium Score Regression (LDSC), drawing on publicly available large-scale Genome-Wide Association Study data for these traits. In addition, PGS were constructed using the same data and applied to two target cohorts for out-of-sample prediction of migraine and its subtypes. These were a depression-enriched cohort (*Australian Genetics of Depression Study*; N=12,601), and an unselected population cohort (*QSkin Study of Sun and Health*; N=16,532). Migraine subtypes were defined according to standard criteria and comprised a broad migraine without aura phenotype and three nested subtypes: migraine with aura, and chronic migraine with and without aura.

We found significant genetic correlations between migraine and depression (*r_g_*=0.29), as well as between migraine and anxiety (*r_g_*=0.32). Across both cohorts, Migraine (*OR*≈1.35) and Depression PGS (*OR*≈1.12) were significantly associated with all measures of migraine and its subtypes. Depression PGS remained significantly associated with all non-chronic migraine subtypes after controlling for migraine and anxiety PGS, suggesting an independent contribution of depression genetic risk on migraine. Anxiety PGS were significantly associated with all non-chronic migraine subtypes (*OR*≈1.09). However, these associations did not persist after adjustment for migraine and/or depression PGS. These results provide insight into the genetic relationships between migraine and its subtypes with depression and anxiety.

---

Migraine is a neurological disorder affecting ∼15% of the global population and ranks as the third leading cause of years lived with disability (YLDs) worldwide, within the category of headache disorders^1^. Characterized by severe and recurring headaches, typically one-sided and often accompanied by nausea and sensitivity to light or sound, migraine manifests in distinct subtypes, such as with or without aura (i.e., visual or sensory disturbances experienced during the attack)^2^. As opposed to episodic migraine, chronic migraine represents a more severe form, characterized by experiencing migraine symptoms on more than 15 days per month, with a population prevalence of approximately 2%^2^. Genetics plays a substantial role in migraine risk, with a heritability of ∼55%, and Genome-Wide Association Studies (GWAS) to date have identified 123 risk loci (i.e., genomic regions) associated with migraine^3^.

Migraine often co-occurs with depression and anxiety, and the disorders share common symptoms, including sleep problems and appetite changes, a higher occurrence in women, and similar prevalence estimates^4–8^. There is evidence supporting causal pathways between these traits; both anxiety and depression double the risk of experiencing migraines, and migraine likewise increases the risk of being diagnosed with anxiety or depression^6,9–11^. This bidirectional pattern is seen specifically in migraine and not in other headache disorders^4,6^.

Previous research suggests that depression and anxiety disorders tend to be more strongly associated with migraine with aura compared to migraine without aura^10,12,13^. This is particularly intriguing, considering that patients who suffer migraine without aura typically experience more severe pain and more frequent attacks^4,13,14^. In addition, individuals with chronic migraines are twice as likely to suffer from depression compared with individuals that experience non-chronic migraine^5^. Additionally, the risk of progressing from episodic to chronic migraine increases with depression severity^5,15,16^. While associations are less clear for anxiety, prior research suggested that chronic migraine shows the strongest association with anxiety, compared to different migraine subtypes or other headache disorders^5,9–11,17^.

Prior research supports a genetic overlap between migraine, depression and anxiety^18–22^. Genetic correlation studies based on twin models or GWAS data consistently show significant genetic correlations between depression and migraine (r_g_ ≈ 0.25-0.33)^13,18,22–25^, as well as between anxiety and migraine (r_g_ ≈ 0.3)^18,24^. Migraine co-occurring with depression may represent a genetically distinct form of migraine compared to ‘pure’ migraine, as several studies have identified differences in heritability patterns between the two phenotypes^18–21^. In particular, patients with comorbid migraine and anxious depression tend to show greater genetic similarity to individuals with depression than to those with pure migraine^18^. In addition, many migraine-associated genetic variants are also implicated in depression^18,20,21,23–26^. However, most of these studies are based on prior underpowered GWAS data. In addition, few studies have examined the relationship between depression and migraine subtypes. For example, migraine with aura shows a stronger genetic association with depression than migraine without aura, supporting epidemiological findings^13^. Beyond this, the genetic relationships between depression, anxiety, and specific migraine subtypes remain unexplored.

This study aims to investigate the genetic relationship between migraine and its subtypes (migraine with and without aura; chronic migraine with and without aura) with depression and anxiety. The availability of recent, well-powered GWAS of migraine provides a unique opportunity to investigate the shared genetic architecture of migraine with depression and anxiety^3^, which we examined using genetic correlations. Given that existing GWAS results for specific migraine subtypes are still underpowered, genetic analyses for these subtypes using GWAS data alone were not feasible. Therefore, we used the well-powered discovery GWAS to construct polygenic scores (PGS) for depression, anxiety, and migraine. These PGS were then applied for out-of-sample prediction of migraine and its subtypes in two large target cohorts. These were a depression-enriched cohort (*Australian Genetics of Depression Study*; N=12,601), and an unselected population cohort used for ascertainment (*QSkin Study of Sun and Health*; N=16,532)^27,28^. PGS represent a measure of genetic risk for a trait, calculated by summing the effects of common genetic variants weighted by their association with that trait.

## Methods

### Participants

This study used data from two Australian cohorts. 1) The Australian Genetics of Depression Study (AGDS); an ongoing depression cohort in which over 22,000 participants were recruited via Australian government prescription records and through a media campaign^27^. In this cohort, participants completed online questionnaires, including an extensive migraine questionnaire. 2) Queensland Skin Study of Sun and Health Study (QSkin), an unselected population cohort that was invited to participate via a random draw from the electoral roll (voter enrolment is compulsory in Australia) and established to investigate skin cancer progression^28^. Although participants did not complete a detailed migraine questionnaire, they indicated a self-reported medical history of migraine.

### Migraine assessment across cohorts

In AGDS, participants were asked, “Have you ever had migraine or recurrent attacks of headaches?” with response options of “Yes” or “No”. Individuals who respond affirmatively to the self-report proceeded to complete a full migraine questionnaire containing 30 items (Supplementary Table 1). Answers to this questionnaire were used to define migraine subtypes. In QSkin, participants were asked, “Have you ever been diagnosed with, experienced, or been treated for any of the following conditions (cross all that apply): Migraine”.

### Migraine subtype classifications in AGDS

We classified migraine into four subtypes based on established diagnostic criteria (*Figure 1, Supplementary Table 1*)^2^:

1. Migraine without aura
2. Migraine with aura
3. Chronic migraine without aura
4. Chronic migraine with aura.

**Figure 1.**
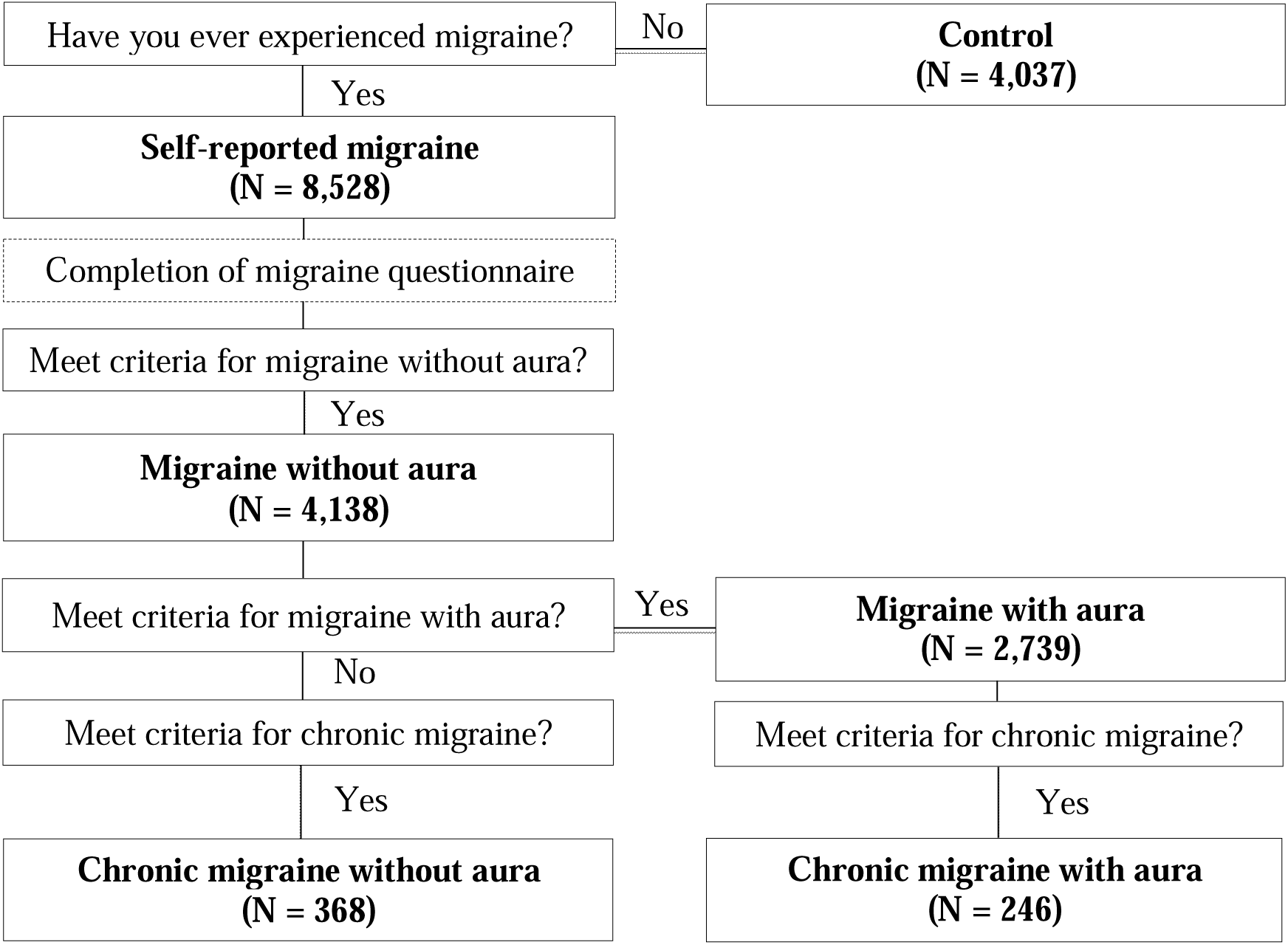
Overview of the Migraine Subtypes Classification Process in AGDS. *Note.* The specific questionnaire items and diagnostic criteria used for classification are provided in Supplementary Table S1. Migraine with aura and chronic migraine subtypes are nested within migraine without aura phenotype.

According to diagnostic criteria^2^, migraine with aura and chronic migraine are additional features that can occur in individuals who meet criteria for migraine without aura. Therefore, migraine with aura, and chronic migraine with and without aura subtypes were nested within migraine without aura, resulting in overlapping samples (*Figure 1, Supplementary Table 1*). Migraine with aura is a migraine-type headache accompanied by transient neurological symptoms (e.g., visual, sensory, or speech disturbances) that precede or accompany an attack^2^. Migraine without aura is a migraine-type headache that occurs in the absence of such symptoms^2^. Chronic migraine is a high-frequency form of migraine, defined by headache occurring on ≥15 days per month, and may occur with or without aura^2^.

The 30-item migraine questionnaire in AGDS was designed to align with the diagnostic criteria for migraine in the second edition of the International Classification of Headache Disorders (ICHD-II)^29^. Here, we used items from this questionnaire to classify migraine and its subtypes following the more recent ICHD-III diagnostic criteria^2^. The instrument was based on the third edition of the deCODE Migraine Questionnaire (DMQ3)^30^, which was explicitly developed for genetic studies and formally validated against a physician-conducted (semi-structured) interview as a gold standard. In the original validation^30^, DMQ3 showed very high sensitivity and specificity for migraine overall, with good agreement for both migraine with aura and migraine without aura.

### GWAS summary statistics selection

Publicly available GWAS summary statistics for depression^31^, anxiety^32^, and migraine^3^, representing the largest available of their kind at time of analysis. Migraine GWAS summary statistics were obtained from the International Headache Genetics Consortium (IHGC). The anxiety GWAS summary statistics were obtained from Purves et al. (2020), which represented the largest available GWAS dataset for generalized anxiety disorder. For depression, a leave-one-out version of the GWAS meta-analysis by Howard et al. (2019)^31^ was used to avoid sample overlap with target AGDS genotype data, which is essential to maintain the validity of polygenic risk scoring.

### Genetic correlation analyses

Linkage Disequilibrium Score Regression (LDSC; release 13/02/2015)^33^ was applied to estimate genetic correlations between migraine and depression, as well as between migraine and anxiety. These analyses used the GWAS summary statistics described above. European LD scores derived from the 1000 Genomes Project were used in the LDSC analysis^34^ and Bonferroni correction was applied for 3 tests to account for multiple testing. Statistical significance was defined as Bonferroni-adjusted *p*<0.05.

### Construction of Polygenic Scores

Polygenic weights were constructed in SBayesR in GCTB (v. 2.03beta)^35^ using the GWAS summary statistics described above, and Polygenic scores were calculated with PLINK (v1.9)^36^ (*Supplementary Material. Polygenic scores construction*). Details on genotyping and genotype data quality control are previously published for AGDS^37^ and QSkin^28^.

### Regression Analyses

Multiple logistic regression analyses were conducted in both cohorts to determine the association between PGS for depression, anxiety and migraine with each of the case/control outcomes: 1. self-reported migraine (AGDS and QSkin), 2. migraine without aura (AGDS), 3. migraine with aura (AGDS), 4. chronic migraine without aura (AGDS) and 5. chronic migraine with aura (AGDS)(*Figure 2*). For each of the 5 case/control outcomes, seven regressions were conducted, with varying combinations of migraine PGS, depression PGS and anxiety PGS as predictor variables (*Figure 2*):

1. *Single PGS model*: each PGS alone (3 regressions);
2. *Pairwise PGS model*: pairwise combinations between the PGS (3 regressions);
3. *Full PGS model*: all three PGS (1 regression).

**Figure 2.**
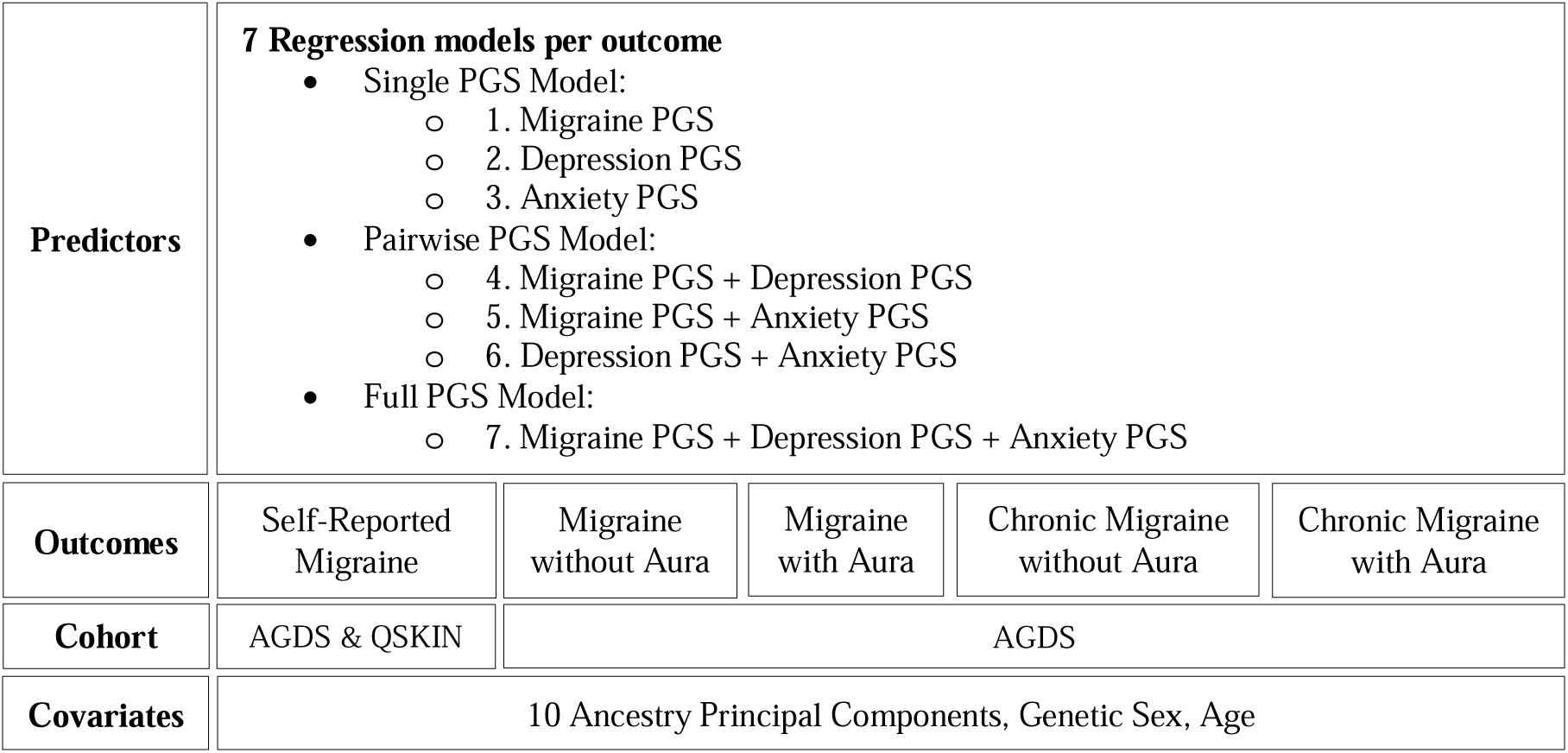
Overview of the Multiple Logistic Regression Analyses.

In both cohorts, only non-related participants were retained (Identity By Descent<0.2 as estimated in PLINK(v1.9))^38^. For each outcome, controls consisted of all individuals who self-reported never experiencing migraine. The PGS were standardized within each cohort, so that effect sizes from logistic regression are reported as odds ratios (ORs) per one standard deviation increase in PGS. To correct for multiple testing, Bonferroni correction was applied separately in each cohort: 49 tests in AGDS, and 7 tests in QSkin. Statistical significance was defined as Bonferroni-adjusted *p*<0.05. Each regression included the following covariates: 10 ancestry principal components, sex (inferred from genotype data), and age. Assumptions of the logistic regression model were evaluated (*Supplementary Material, Model Checking*). All statistical analyses were conducted in R(version 4.3.3) and RStudio(version 2023.12.1)^39^. A sensitivity analysis using alternative controls for chronic migraine was performed, where individuals with chronic migraine were compared with a control group comprising controls and individuals with non-chronic migraine (*Supplementary Material, Alternative Non-Chronic Control Group*).

### Power analyses

Power calculations were conducted using the *pwr* package in R^40^. The parameters required for this package include the effect size (*F²*), total sample size, number of predictors (*k*=15; depression PGS, anxiety PGS, migraine PGS, sex, age, 10 PCs), and a significance level set at 0.05. The effect size F² was calculated from a target OR of 1.1, which was converted to Cohen’s *d* and then to *F²* using standard methods^41,42^. Power>0.8 was considered adequate.

### Use of Artificial Intelligence Tools

Generative AI (ChatGPT, OpenAI) was used to assist with language editing. The authors take full responsibility for the content of the manuscript.

## Results

### Genetic Correlation

LDSC analyses revealed a significant positive genetic correlation between migraine and depression (*r_g_*=0.29, *SE*=0.02, *z*=14.59, *p_adj_=*6.20e-48), as well as between migraine and anxiety (*r_g_*=0.32, *SE*=0.04, *z*=8.68, *p_adj_=*7.62e-18; *Supplementary Table 2*).

### Polygenic Score Analyses

#### Cohort characteristics

The analyses included 12,601 participants from AGDS, the depression-enriched cohort, and 16,532 participants from QSkin, the unselected population cohort (*Table 1*). AGDS consisted of 8,528 individuals who self-reported migraine and 4,073 controls. The migraine subtypes included 4,138 individuals qualifying for migraine without aura, 2,739 for migraine with aura, 368 for chronic migraine without aura, and 246 for chronic migraine with aura. In QSkin, 2,701 individuals self-reported migraine, and 13,831 were classified as controls. Compared to QSkin, AGDS had higher proportions of self-reported depression (94.7% vs. 16.6%), females (75.7% vs. 54.9%) and self-reported migraine (67.7% vs. 16.3%), consistent with the higher prevalence of depression in females and the higher prevalence of migraine among people with depression. QSkin participants were on average 16 years older than those in AGDS. Conducting analyses in these two cohorts with differing demographic and clinical profiles enables assessment of associations across both clinically enriched and population-based samples.

**Table 1.** Summary of Participant Characteristics Included in the Analyses.

|  | AGDS | QSkin |
| --- | --- | --- |
| **Cohort type** | Depression-enriched | Unselected population |
| **Total number of participants** | 12,601 | 16,532 |
| **Self-reported migraine (N)** | 8,528 | 2,701 |
| **Migraine with aura (N)** | 2,739 | - |
| **Migraine without aura (N)** | 4,138 | - |
| **Chronic migraine with aura (N)** | 246 | - |
| **Chronic migraine without aura (N)** | 368 | - |
| **Self-reported depression (%)** | 94.71 | 16.56 |
| **Female (%)** | 75.73 | 54.88 |
| **Age mean in years (SD)** | 44.23 (15.25) | 60.91 (8.92) |

#### Results from the AGDS cohort

##### Power analyses

Models for self-reported migraine, and migraine with and without aura all had power>0.8 to detect an OR≥1.1 at a significance level of 0.05. However, chronic migraine analyses had low power to detect an OR≥1.1(power=0.13 for migraine without aura; power=0.10 for migraine with aura)(*Supplementary Table 4*).

##### Migraine PGS

Migraine PGS were significantly associated with higher odds for all migraine phenotypes across all PGS models: self-reported migraine (*OR*=1.29, 95%*CI*[1.24,1.34], *p_adj_=*1.07e-32, *full model*), migraine without aura (*OR*=1.4, 95%*CI*[1.33,1.47], *p_adj_*=3.64e-41, *full mode*l), migraine with aura (*OR*=1.41, 95%*CI*[1.34,1.49], *p_adj_*= 3.06e-35, *full model*), chronic migraine without aura (*OR*=1.28, 95%*CI*[1.15,1.43], *p_adj_*=2.28e-4, *full model*), and chronic migraine with aura (*OR*=1.28, 95%*CI*[1.13,1.46], *p_adj_*=7.23e-3, *full model*)(*Figure 3, Supplementary Table 3)*.

**Figure 3.**
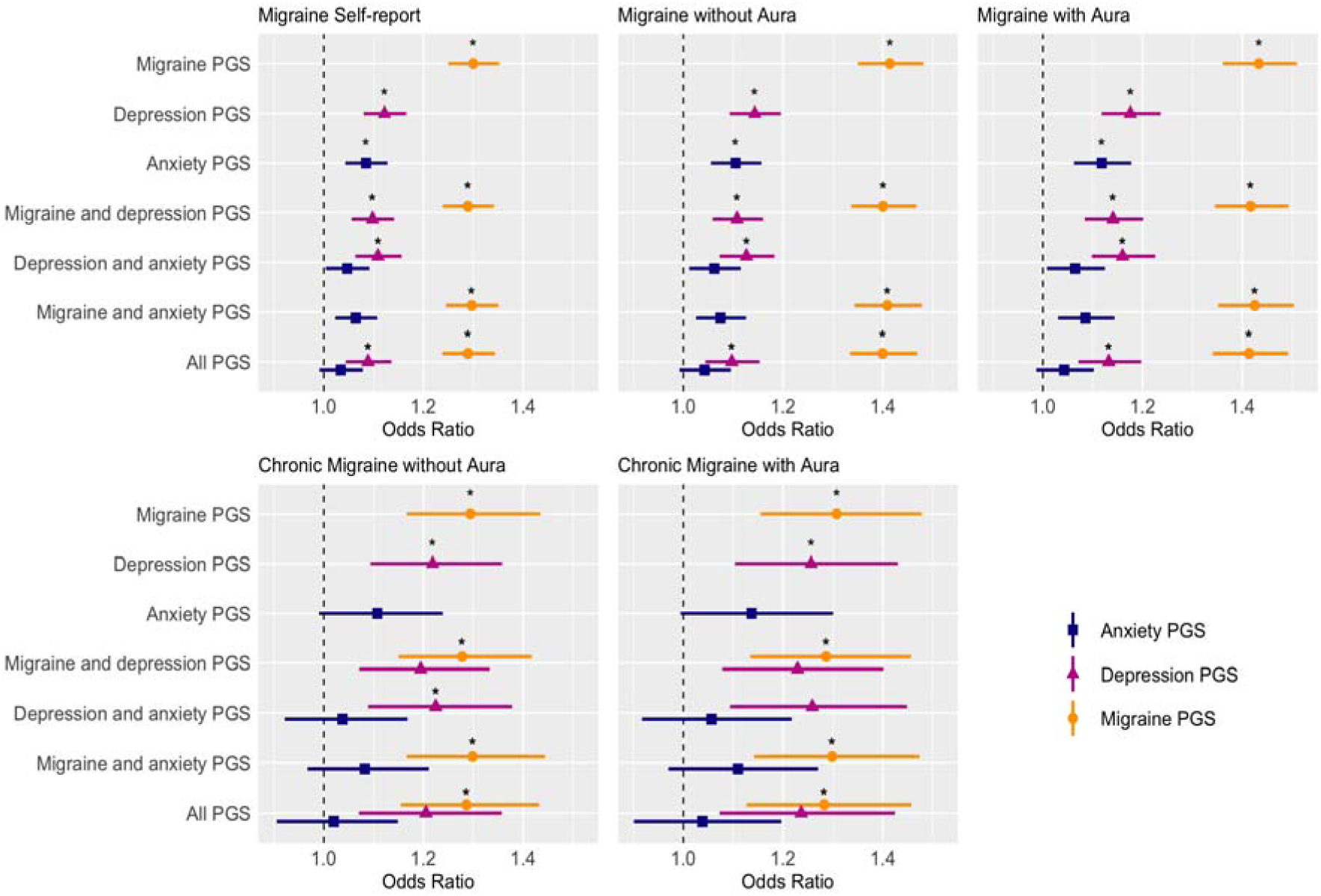
Forest Plots showing Odds-Ratio for Polygenic Score (PGS) Associations Across Migraine Phenotypes in the AGDS Cohort. Note: Central points represent Odds-Ratio estimates, and horizontal lines indicate the corresponding 95% confidence intervals before Bonferroni correction. Stars denote associations that remained significant after Bonferroni correction. Colors and shapes represent each trait (orange/circle = migraine PGS; pink/triangle = depression PGS; blue/square = anxiety PGS). Analyses were sufficiently powered (power > 0.80 to detect an OR ≥1.1 at a significance level of 0.05) for migraine self-report, migraine with aura, and migraine without aura, but was low for chronic migraine without aura (power = 0.13) and chronic migraine with aura (power = 0.10).

##### Depression PGS

In the single PGS models, depression PGS was significantly associated with higher odds for all migraine phenotypes; self-reported migraine (*OR*=1.12, 95%*CI*[1.08,1.17], *p_adj_*=1.68e-07), migraine without aura (*OR*=1.14, 95%*CI*[1.09,1.2], *p_adj_*=2.51e-07), migraine with aura (*OR*=1.18, 95%*CI*[1.12,1.24], *p_adj_*=1.43e-08), chronic migraine without aura (*OR*=1.22, 95%*CI*[1.09,1.36], *p_adj_*=1.69e-2), and chronic migraine with aura (*OR*=1.26, 95%*CI*[1.1,1.43], *p_adj_*=2.78e-2)(*Figure 3, Supplementary Table 3).* When adjusting for migraine PGS, the associations between depression PGS and all non-chronic migraine phenotypes were slightly attenuated, but remained significant (self-reported migraine, *OR*=1.09, 95%*CI*[1.04,1.14]; migraine without aura, *OR*=1.1, 95%*CI*[1.04,1.15]; migraine with aura, *OR*=1.13,95%CI[1.07,1.2]; *full model*) (*Figure 3, Supplementary Table 3*). Associations between depression PGS and chronic migraine subtypes did not survive multiple testing correction after adjusting for migraine PGS, although a positive trend remained (chronic migraine without aura, *OR*=1.2, 95%*CI*[1.07,1.36], *p_adj_*=9.85e-2; chronic migraine with aura, *OR*=1.23, 95%*CI*[1.07,1.42], *p_adj_*=1.62e-1; *full model*) (*Figure 3, Supplementary Table 3)*, reflecting the low power of the chronic migraine analyses (*Supplementary Table 6*). When adjusting for anxiety PGS, the associations between depression PGS and migraine phenotypes remained predominantly unchanged compared to the single PGS model, except for chronic migraine with aura that no longer reached statistical significance, reflecting the low power of the chronic migraine analyses (*Supplementary Table 6*).

##### Anxiety PGS

In the single PGS models, anxiety PGS was significantly associated with higher odds for all non-chronic migraine phenotypes; self-reported migraine (*OR*=1.08, 95%*CI*[1.04,1.13], *p_adj_*=2.107e-3), migraine without aura (*OR*=1.1, 95%*CI*[1.06,1.16], *p_adj_*=9.849e-4), migraine with aura (*OR*= 1.12, 95%*CI*[1.06,1.18], *p_adj_*=9.604e-4). In the pairwise and full models adjusting for migraine and/or depression PGS, anxiety PGS showed weaker and no longer significant associations (*Figure 3, Supplementary Table 3*). Anxiety PGS showed a positive non-significant association with chronic migraine phenotypes in the single PGS model (chronic migraine without aura: *OR*=1.11, 95%*CI*[0.99,1.24], *p_adj_*=1; chronic migraine with aura: *OR*=1.14, 95%*CI*[0.99,1.3], *p_adj_*=1) (*Figure 3, Supplementary Table 3)*. This trend was attenuated in models adjusting for migraine and/or depression PGS (chronic migraine without aura: *OR*=1.02, 95%*CI*[0.91,1.14], *p_adj_*=1; chronic migraine with aura: *OR*=1.14, 95%*CI*[0.9,1.2], *p_adj_*=1; *full model*) (*Figure 3, Supplementary Table 3)*. The chronic migraine analyses are associated with low power, however (*Supplementary Table 6*).

#### Results from the QSkin cohort

Associations between PGS and self-reported migraine history in QSkin were consistent with those observed in AGDS. Migraine subtype information was not available in QSkin.

##### Migraine PGS

Migraine PGS were significantly associated with higher odds of self-reported migraine across all models (*OR*=1.41, 95%*CI*[1.36,1.47], *p_adj_*=1.97e-59, *full model*)(*Figure 4, Supplementary Table 5*). The effect sizes remained very similar across all models, and all had power > 0.8 to detect an OR ≥1.1 at a significance level of 0.05 (*Supplementary Table 4*).

**Figure 4.**
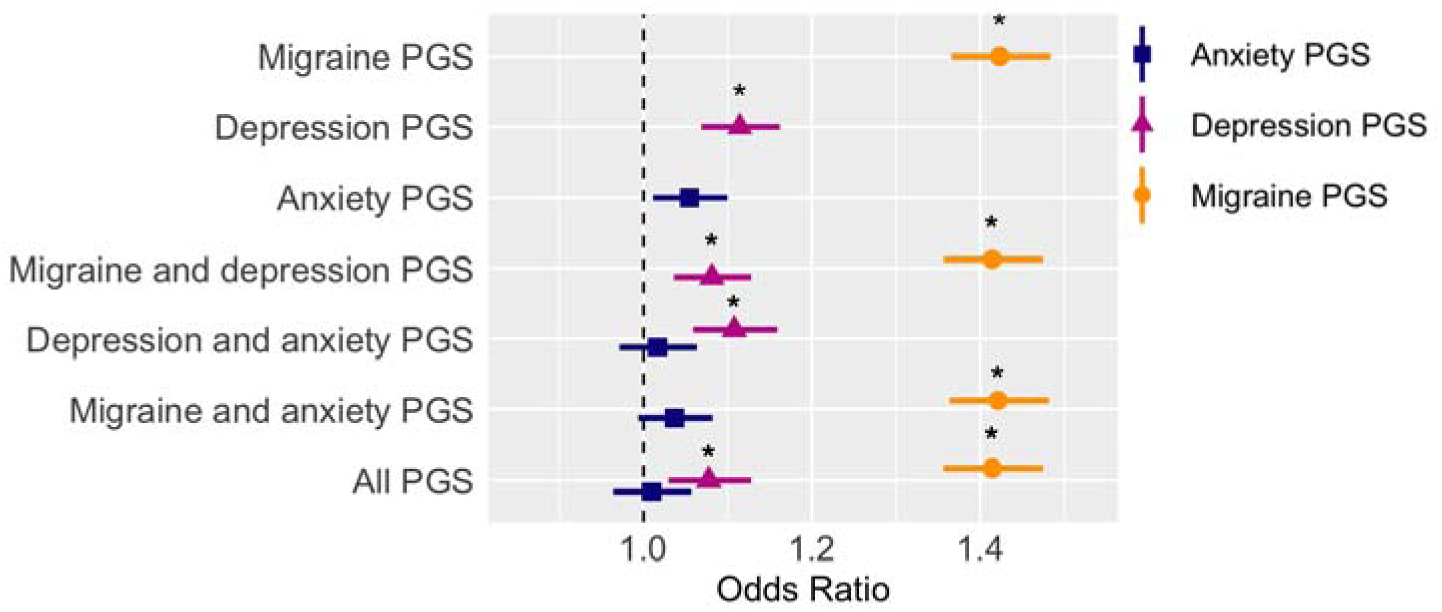
Forest Plot Showing Odds-Ratio for Polygenic Score (PGS) Associations with Self-Reported Migraine in the QSkin Cohort. Note: Central points represent Odds-Ratio estimates, and horizontal lines indicate the corresponding 95% confidence intervals before Bonferroni correction. Stars denote associations that remained significant after Bonferroni correction. Colors and shapes represent each trait (orange/circle = migraine PGS; pink/triangle = depression PGS; blue/square = anxiety PGS). Analyses were sufficiently powered (power > 0.80 to detect an OR ≥1.1 at a significance level of 0.05)

##### Depression PGS

Depression PGS were significantly associated with higher odds of reporting migraine across all models, and effect sizes were the highest in the single PGS model (*OR*=1.11, 95%*CI*[1.07,1.16], *p_adj_*=2.84e-06). When controlling for migraine PGS, the association between depression PGS and self-reported migraine was slightly attenuated in both the pairwise and full models (*OR*=1.08, 95%*CI*[1.03,1.13], *p_adj_*=8.74e-3, *full model*)*(Figure 4, Supplementary Table 5*).

##### Anxiety PGS

Although associations did not survive multiple testing correction, anxiety PGS were nominally associated with higher odds of reporting migraine (single PGS model; *OR*=1.05, 95%*CI*[1.01,1.1]). This trend remained after adjusting for migraine PGS (pairwise model; *OR*=1.04, 95%*CI*[1.00,1.08]). However, it was no longer observed when depression PGS was included in the models (*OR*=1.01, 95%*CI*[0.96,1.06]; *full model*)(*Figure 4, Supplementary Table 5*). This pattern is consistent with findings observed in the AGDS cohort.

## Discussion

Migraine, depression, and anxiety are highly disabling disorders that share clinical features and often co-occur. This study investigated the genetic relationship between these traits using genetic correlation and polygenic score (PGS) analyses. We observed significant genetic correlations between migraine and depression, as well as between migraine and anxiety. Polygenic scores (PGS) for the three traits were applied to two target cohorts (a depression-enriched and an unselected population used for ascertainment) to assess their association with migraine, as well as migraine subtypes, which were assessed only in the depression-enriched cohort. Migraine PGS were positively associated with migraine and all its subtypes. Depression PGS showed a positive and independent association with migraine, beyond the genetic contributions of migraine and anxiety. Anxiety PGS were also positively associated with migraine, however, this effect was no longer present after controlling for depression PGS.

Using the largest available GWAS for these traits at the time^3,31,32^, we found a significant genetic correlation between migraine and depression (r_g_=0.29, se=0.02), as well as between migraine and anxiety (r_g_=0.32, se=0.04). These estimates are consistent with prior genetic correlation estimates with both depression^13,18,22–25,43^ and anxiety^18,24^. Recent anxiety GWAS have reported slightly lower genetic correlations with migraine (∼0.2), likely reflecting broader and more heterogeneous anxiety phenotypes relative to the generalized anxiety disorder-focused GWAS used here^44^. These results suggest shared genetic influences and potential pleiotropy between these traits. Pleiotropic effects between migraine and depression have been reported previously. Yang et al. (2018) identified three SNPs associated with both migraine and depression, while Bahrami et al. (2022) reported 37 genes implicated in both conditions^23,45^. Evidence for specific shared loci between migraine and anxiety remains limited, largely due to the relatively low power of existing anxiety GWAS.

Migraine PGS were positively associated with self-reported migraine across both cohorts, and with all migraine subtypes that were examined in the depression-enriched cohort. These associations showed comparable effect sizes across migraine subtypes and cohorts. This is consistent with current migraine GWAS capturing genetic variation to overall migraine. GWAS findings for migraine subtypes currently remain underpowered, limiting the identification of subtype-specific loci^3^. Prior evidence suggests that the genetic architecture of migraine with and without aura is highly similar, particularly at the level of common genetic variants^46^. For chronic migraine, prior work supports our results showing that migraine PGS exhibited comparable effect sizes with chronic migraine and overall migraine^47^. Other evidence suggests that migraine chronification shows limited familial aggregation and is driven predominantly by environmental factors, including medication overuse, stress, and depression^16^.

Depression PGS were also positively associated with migraine, and the associations remained after adjusting for both migraine and anxiety PGS. This suggests that depression PGS confers a distinct and independent genetic contribution to migraine, over and above the genetic influences of migraine and anxiety. The effect sizes were highly comparable across migraine subtypes and cohorts, suggesting that the results are relevant to the general population. Chronic migraine subtypes showed nominal associations that did not survive correction for multiple testing; however, this most likely reflects their lower statistical power, as the overall pattern of effect sizes remained highly similar across subtypes. This homogeneity in effect sizes contrasts with prior research that reported a stronger genetic association between depression and migraine with aura^13^. One possible explanation is the use of nested migraine subtypes in our analyses based on the ICHD-III classification introduced in 2017, whereas earlier studies relied on mutually exclusive ICHD-II categories. However, recent evidence indicating a highly similar genetic architecture between migraine with and without aura also supports our findings^46^.

Anxiety PGS were positively associated with migraine and its subtypes. In the depression-enriched cohort, anxiety PGS alone was positively associated with migraine with and without aura. However, these associations were substantially attenuated after adjustment for migraine and/or depression PGS and no longer remained significant. The pattern of effect sizes was highly comparable across migraine subtypes and cohorts, suggesting that the findings are relevant to the general population. This is likely due to the lower statistical power of the anxiety GWAS relative to the depression GWAS, combined with the high genetic overlap between the two traits (rg ≈ 0.9), which limits the reliable detection of anxiety-specific effects and does not necessarily indicate a weaker underlying biological relationship between anxiety and migraine^48^. Controlling for migraine PGS also modestly attenuated the effect of anxiety PGS, although to a lesser extent than adjustment for depression PGS. This pattern may reflect shared genetic liability between migraine and anxiety, but could likewise be influenced by the comparatively lower statistical power of the anxiety GWAS.

This study should be interpreted considering several limitations. First, the findings are correlational and therefore do not establish causality. Second, the cohorts and GWAS data used are derived from individuals of European ancestry. Third, migraine was assessed via self-report, which may introduce misclassification. Moreover, migraine subtypes were examined exclusively in the depression-enriched cohort, where these measures were available, potentially introducing selection bias. Finally, the chronic migraine analyses were underpowered due to small sample size, limiting interpretability.

Moving forward, larger and better-powered GWAS for anxiety and migraine subtypes, particularly for the presence of aura, are needed to better capture trait- and subtype-specific genetic variation. In addition, investigating migraine subtypes, particularly chronic migraine, within a larger cohort not selected for depression, may help disentangle the specific genetic relationships with more precision. While anxiety is most often studied in the context of co-occurring depression, examining anxiety independently is necessary to clarify its unique genetic contributions to migraine. Causal inference approaches, including Mendelian randomization could be incorporated in future analyses, especially with the availability of larger and better-powered GWAS.

## Data Availability

The source data and scripts used for all analyses will be made available in a public repository. Individual-level genotype data cannot be shared.

## Author Contributions

C.D.: Data curation, Formal analysis, Investigation, Methodology, Software, Visualization, Writing - original draft. J.T.: Conceptualization, Data curation, Methodology, Supervision, Writing - review & editing. B.M.: Conceptualization, Data curation, Funding Acquisition, Resources, Methodology, Supervision, Writing - review & editing. D.N.: Methodology, Writing - review & editing. Acquisition. N.M.: Writing - review & editing. Other contributors: Cohort creation & Funding

## Acknowledgements

We are indebted to the participants for giving their time to contribute to this study. We thank all the people who helped in the conception, implementation, beta testing, media campaign, and data cleaning.

