## Supplementary Material for "Exploring the Genetic Relationship Between Migraine Subtypes, Depression And Anxiety Using Polygenic Scores"

### Supplementary Methods

#### Polygenic Scores construction

Polygenic weights were constructed using SBayesR in GCTB (v. 2.03beta)[1] and the provided linkage disequilibrium eigen-decomposition data from UK Biobank participants of European ancestry (7 million imputed SNPs). Analyses were run with a chain length of 25,000 with a burn-in of 5000, and the remaining default settings. These polygenic weights were then used to calculate polygenic scores (PGS) using genotype data form each of the two target cohorts using PLINK (v1.9)[2], summing the number of effect alleles weighted by their posterior mean effect estimates (using the flag --score).

#### Model checks

Logistic regression model assumptions were evaluated using multiple diagnostic approaches. Simulation-based residual analyses were conducted using the DHARMa package in R (version 4.3.3)[3,4], which generates scaled residuals by comparing observed data against simulated values from the fitted model. Uniformity of residuals was assessed using the Kolmogorov-Smirnov (KS) test, and dispersion was evaluated using a non-parametric test comparing the standard deviation of observed versus simulated residuals. For both tests, $p < 0.05$ indicated potential model misspecification. No significant deviations from uniformity or overdispersion were detected, indicating adequate model fit. Multicollinearity among predictors was assessed using variance inflation factors (VIF), with all values below 5, suggesting no significant collinearity. Visual inspection of residual plots did not reveal influential outliers or systematic patterns indicative of model misspecification.

#### Alternative Non-Chronic Control Group

We tested a secondary control group consisting of everyone who did not meet the criteria for chronic migraine (self-reported never experiencing migraine + non-chronic migraine patients). This secondary set of analyses aimed to identify if there is a difference in genetic risk between chronic migraine compared to non-chronic migraine (*Supplementary Figure 1*).

#### **Supplementary Results**

*Alternative Non-Chronic Control Group*

For chronic migraine, we performed additional analyses using an alternative control group that combined the original controls (i.e., participants who self-reported not experiencing migraine) with all participants who reported experiencing migraine but did not meet criteria for chronic migraine. Although no significant associations were observed for any PGS, which may be due to the low power associated with the chronic migraine analyses, there were patterns of non-significant trends. Depression PGS showed the strongest association with chronic migraine with and without aura (chronic migraine without aura, *OR*=1.14, 95%*CI*[1.03,1.33]; chronic migraine with aura, *OR*=1.17, 95%*CI*[1.02,1.34]; *full model*, *Supplementary Figure 1*). Associations between migraine PGS and chronic migraine also showed a positive trend (chronic migraine without aura, *OR*=1.1, 95%*CI*[0.99,1.22]; chronic migraine with aura, *OR*=1.10, 95%*CI*[0.97,1.25]; *full model, Supplementary Figure 1*).

**Supplementary Figure 1**

*Forest plots showing Odds-Ratio for Polygenic Score (PGS) associations across chronic migraine phenotypes in the AGDS cohort with an alternative non-chronic control group consisting of everyone not meeting criteria for chronic migraine.*

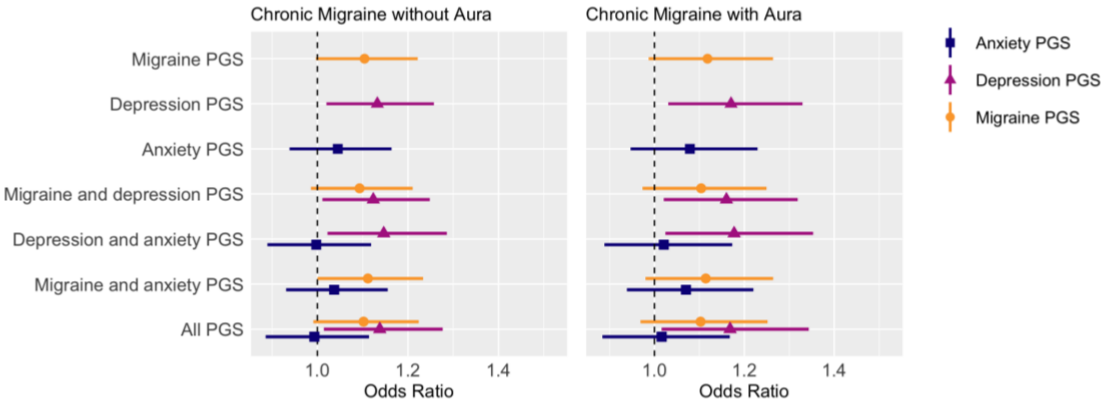


Note: Central points represent *Odds-Ratio* estimates, and horizontal lines indicate the corresponding 95% confidence intervals before Bonferroni correction. Stars denote associations that remained significant after Bonferroni correction. Colors and shapes represent each trait (orange/circle = migraine PGS; pink/triangle = depression PGS; blue/square = anxiety PGS). Statistical power was low for chronic migraine with aura (0.10) and without aura (0.13) to detect OR ≥ 1.1 at a significance level of 0.05.
